# Mucin 5AC is a sensitive surface marker for sessile serrated lesions: results from a systematic review and meta-analysis

**DOI:** 10.1101/2024.02.11.24302644

**Authors:** Kevin Liu, Moniyka Sachar, Violeta Popov, Ziheng Pei, Giulio Quarta

## Abstract

Sessile serrated lesions (SSLs) are a class of colon polyps which are challenging to detect through current screening methods but are highly associated with colon cancer. We reasoned that a biomarker sensitive for SSLs would be clinically useful to improve detection. Recent endoscopic and histopathologic studies suggest that SSLs are associated with alterations in intestinal mucin expression but the frequency with which this occurs is not known. We performed a meta-analysis of available pathologic studies comparing mucin expression on SSLs to normal colonic mucosa, tubular adenomas (TAs), villous adenomas (VAs), traditional serrated adenomas (TSAs), and hyperplastic polyps (HPs). We searched Medline, Pubmed, and Embase and found 440 publications in this topic, and 18 total studies met inclusion. We found that MUC5AC expression was more common in SSLs compared to normal colonic mucosa (OR=82.9, p<0.01), TAs (OR=11, p<0.01), and TSAs (OR=3.6, p=0.04). We found no difference in MUC5AC expression between SSLs versus HPs (OR=2.1, p=0.09) and no difference in MUC5AC expression between left colon and right colon HPs, with an OR=1.8, p=0.23. We found that MUC5AC expression was found commonly on VAs, SSLs, and TSAs while the frequency on colon cancers declined. MUC5AC is also upregulated in inflammatory bowel disease and in response to intestinal infections. MUC5AC expression highlights the potential of mucins as sensitive biomarkers, though not specific to SSLs. Further research into the clinical utilization of MUC5AC could enhance SSL detection.

## Introduction

In the United States and globally, populations have benefitted greatly from colon cancer screening efforts, with fecal immunochemical testing (FIT), colonoscopy, and computed tomographic colonography the most widely used tests (1). A significant improvement in morbidity and mortality comes from the ability to detect and remove colon polyps (2,3), neoplastic lesions in the colon which are precursors to high-risk dysplasia or cancer. Colon polyps are defined by histopathology as hyperplastic polyps (HPs), tubular adenomas (TAs), villous adenomas (VAs), or sessile serrated lesions (SSLs) and provide prognostic information on an individual risk of cancer (4).

Detecting colon polyps is a major goal of colonoscopy (5), however SSLs in particular have evaded currently available screening methods (6). SSLs are thought to be the most common cause of interval colon cancers; which are cancers diagnosed despite screening (7) and are thought to arise either from an unusually rapid carcinoma sequence or missed lesions not visualized during colonoscopy (7,8). SSLs are typically right-sided lesions, morphologically flat, with histological features of dilated, mucin-filled, serrated crypts (9). SSLs display reduced expression of mismatch repair proteins MLH1 and MLH2 and are associated with mutations in *BRAF* or *KRAS* (10). SSLs were differentiated pathologically from HPs in 2003, and the WHO criteria for the pathological diagnosis of SSLs was updated recently (4,11).

Previous efforts to improve the detection of SSLs in the colon have included methods to examine methylation patterns in DNA (12–15), chromoendoscopy with narrow band imaging (16), color enhancement (17,18), and novel artificial intelligence approaches (19). Despite these efforts, no single modality has gained acceptance due to cost and modest sensitivity (14). We hypothesized that a biomarker akin to the widely available fecal immunochemical testing (FIT), would be a reasonable marker for SSL development. To search for a biomarker of SSLs, we performed a meta-analysis of immunohistochemical studies and found numerous studies examining an altered pattern of mucin expression on the surface of SSLs. Our findings support that the mucin MUC5AC is a sensitive surface marker of SSLs.

Mucins are glycoproteins expressed on the surface of colon epithelium, secreted either by enterocytes or goblet cells (Figure 1) forming a dual layer of protective proteins for epithelial cells (20). The normal colon secretes mucins MUC1, MUC2, and MUC3, which can be membrane-bound or unbound (21,22). However, endoscopists (23) and pathologists (24) have noted abnormalities in the mucin expression pattern on polyps and colon cancers, and this pattern remains a criterion for detection of SSLs. While MUC5AC is typically expressed on surface and pit epithelia of gastric mucosa (25) as well as airway epithelia (26,27), aberrant mucin expression in the colon is thought to arise from the same genetic alterations which cause colon cancer (Figure 1) (23,28).

**Figure 1.**
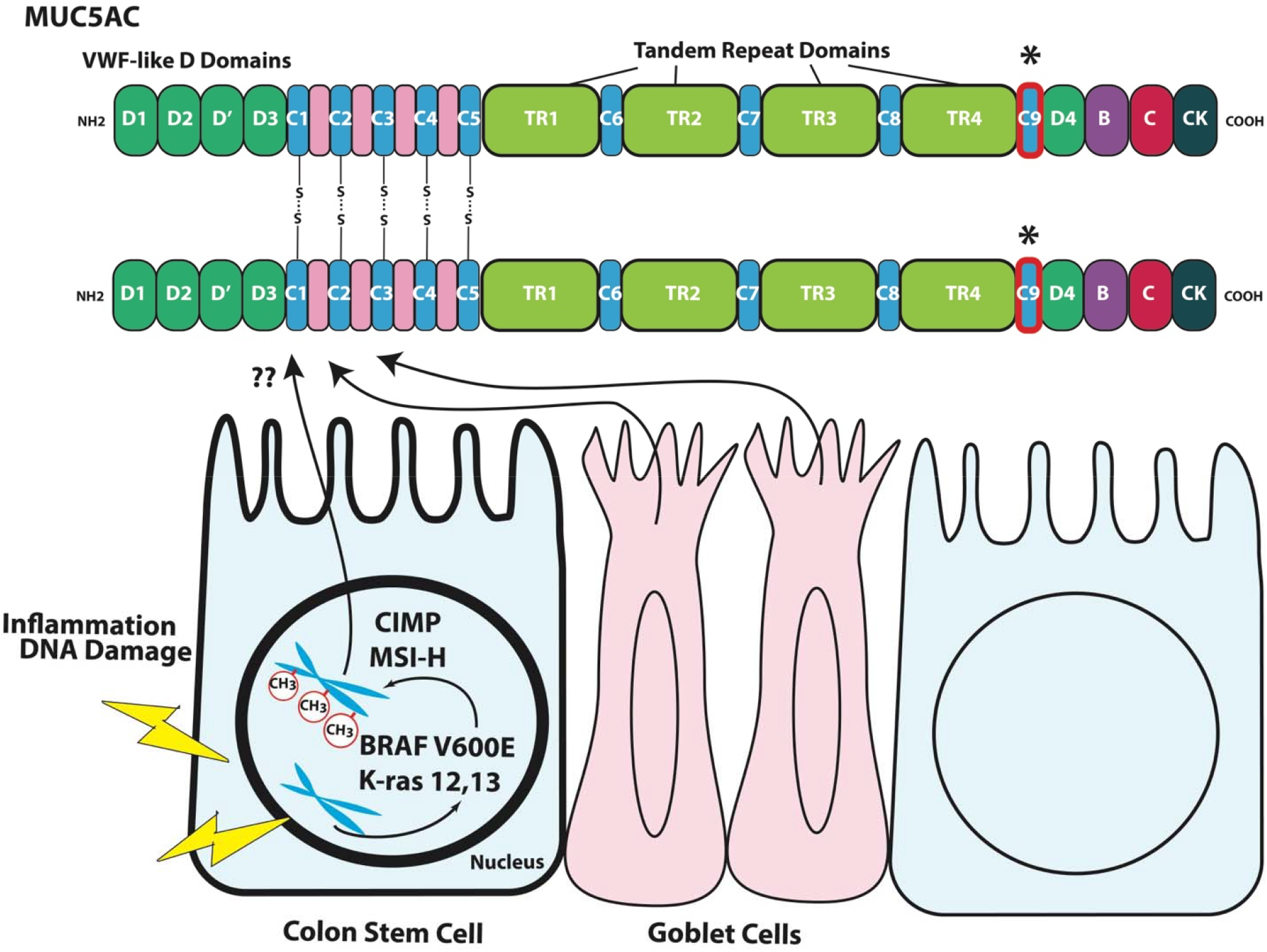
Domain architecture and model of MUC5AC overexpression in sessile serrated carcinogenesis. <u>Top panel.</u> MUC5AC is a polymeric mucin encoded by 9 cysteine rich domains (C1-9) interspersed by nonrepetitive and tandem repeat domains. Cysteine rich domains can form multimers with other mucins (S-S bonds). The 3′-flanking region consists of the cysteine rich domains D4, B, C and cysteine knot (CK). <u>Bottom panel.</u> Inciting mutations in BRAF (V600E) or Kras (Codons 12 or 13) trigger a cascade of aberrant methylation in chromosomal DNA (shown as red circles on DNA) marked by CpG island methylation (CIMP) and microsatellite instability (MSI). One common methylation change is the hypomethylation of MUC5AC promoter region which leads to overexpression and secretion.

In this study, we examined the mucin expression pattern across colon polyps by histopathologic subtype. We find that MUC5AC is a surface marker highly expressed on the surface of SSLs, though it is not specific to serrated pathway lesions.

## Methods

A total of three databases including Embase, Pubmed, and Medline were searched from inception to November 2023. A search strategy using the search criteria “mucin” AND “immunohistochemistry” AND “polyp” to identify studies reporting mucin expression in colonic polyps. Peer reviewed journal articles but not conference abstracts were included in the analysis. Observational retrospective studies and case series were included for full review if they were human studies that reported at least colonic sessile serrated lesion pathology subtypes and the number of colonic polyps associated with mucin positivity. For a study to be included in a MUC5AC expression comparison group, the authors must report mucin expression in polyp subtypes compared. Other reasons for exclusion were if they were written in a language other than English, were conference abstracts, or reported insufficient data to calculate odds ratios. The primary outcome assessed was the expression of mucins on the surface of SSLs compared to normal mucosa, TAs, TSAs, and HPs. Secondary outcomes included the expression of mucins in proximal HPs versus distal HPs. Studies included are listed in Supplementary Table 1 (29–47).

Full texts were reviewed by 2 study authors (KL and MS). Data was abstracted from the full text by one author (either KL or MS) and confirmed by an alternative author (either KL or MS). Conflicts were resolved by discussion and consensus, with a senior author (GQ or VP) serving as the final arbiter if consensus was not achieved.

A random effects meta-analysis was used due to the initial assumption of heterogeneity among the individual studies. Heterogeneity was assessed using Cochran’s Q and I 2 statistics, classified as not important (0 %–40 %), moderate (30 %–60 %), substantial (50 %–90 %), and considerable (75 %–100 %) (Higgins and Green, 2011). All analyses were done using Comprehensive Meta-Analysis Software, version 3.0 (Biostat, Engelwood, NJ, USA).

## Results

Of the 440 citations identified, 18 were included in the final analysis (Fig. 2). Included studies encompassed 3,554 polyps, of which 32-75% came from female patients, with an average age ranging from 57 to 67 years old. Studies included polyp samples from the USA, Japan, Iran, Korea, France, and Turkey (Supplementary Table 1). For MUC1, MUC2, MUC5AC, and MUC6 expression there was insufficient data to perform a meta-regression by age and gender.

**Figure 2.**
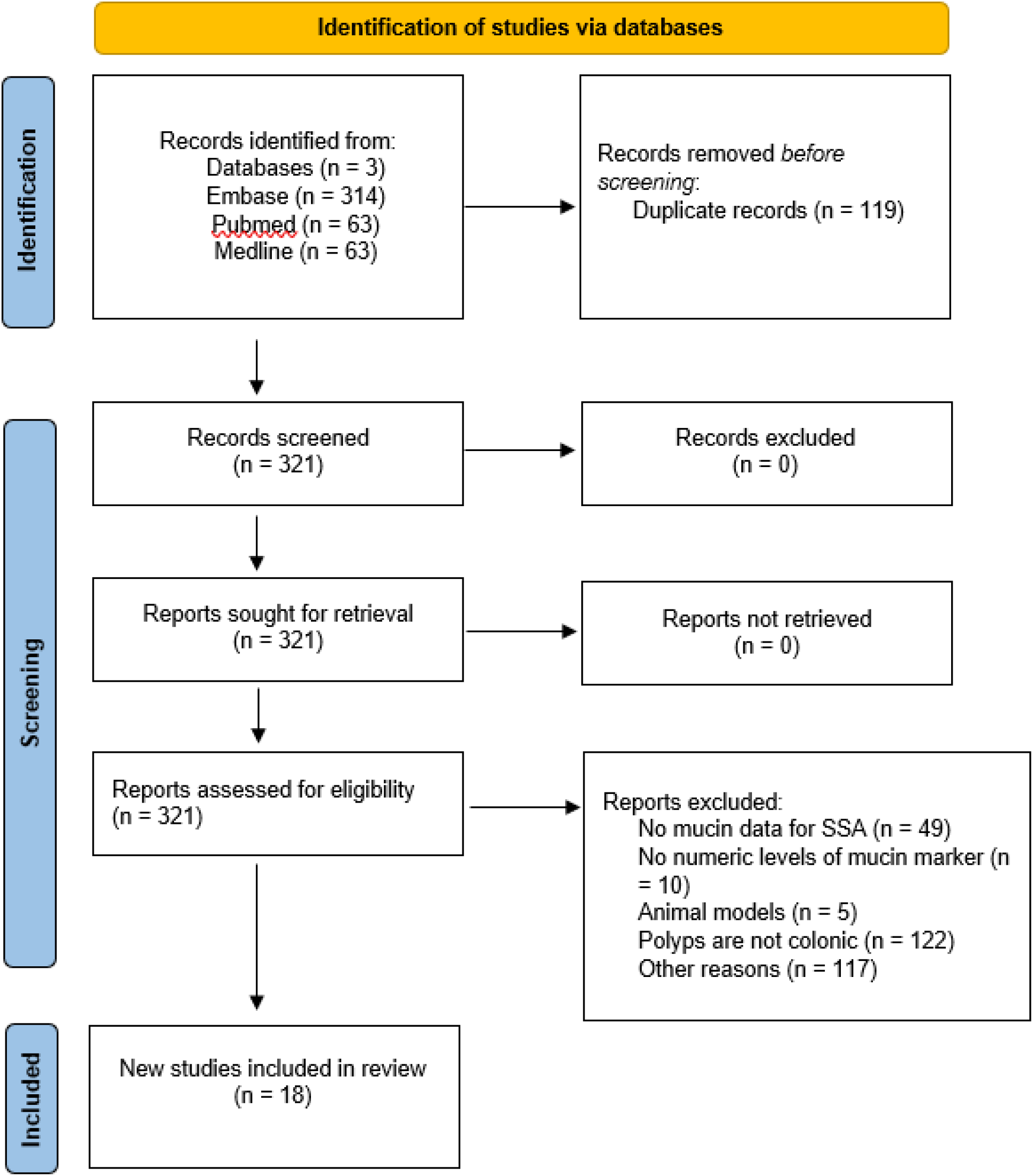
Flowchart of inclusion and exclusion criteria for studies.

**Figure 3.**
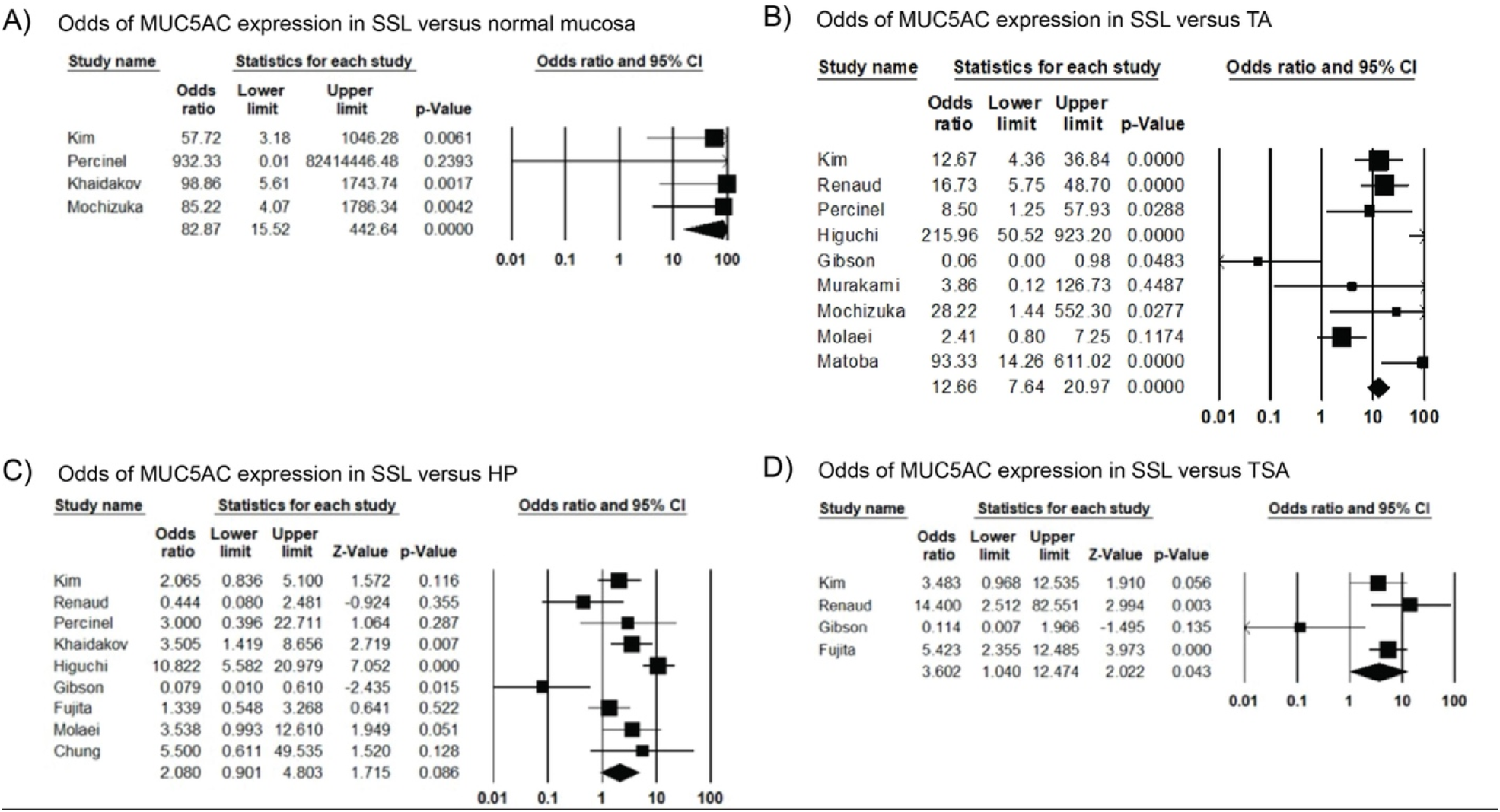
Forest plots of the odds ratios of MUC5AC expression in SSLs versus other histopathologic subtypes. Plots are demonstrating the odds of MUC5AC expression on the surface of SSL versus normal mucosa (A), tubular adenomas (B), hyperplastic polyps (C), and traditional serrated adenomas (D). Studies included in the meta-analysis are shown on the left panel with the cumulative odds ratio and p-values shown on the last line.

Of the 18 studies that were included in the final analysis, 14 reported MUC5AC expression in SSLs (Supplementary Table 2). We found that MUC5AC expression was more common in SSLs compared to normal colonic mucosa, with an OR=82.9, p<0.01 (4 studies, 107 SSLs polyps, 86 normal samples, I2=0%) (Figure 2). This finding is consistent with prior research as MUC5AC is not normally expressed in normal colonic mucosa (21). We found MUC5AC expression in SSLs to be higher than in TA samples, with an OR=11, p<0.01 (9 studies, 412 SSLs polyps, 1221 TA polyps, I2= 82%) and TSA samples, OR=3.6, p=0.04 (4 studies, 287 SSLs polyps, 115 TSA polyps). We found no difference in MUC5AC expression between SSLs versus HP samples, with an OR=2.1, p=0.09 (9 studies, 443 SSLs polyps, 681 HP polyps).

Prior research suggests that a high proportion of proximal HPs show features of SSLs (48). Because proximal HPs and SSLs share such pathologic similarities, we wanted to assess whether proximal HPs would also express increased MUC5AC in comparison to distal HPs. There was no difference in MUC5AC expression between left colon and right colon HP samples, with an OR=1.8, p=0.23 (3 studies, 77 right HP polyps, 151 left HP polyps).

Numerous studies have examined MUC5AC expression in the colon in various pathologic states. MUC5AC is absent from the normal intestine past 12 weeks of gestation (49). Forgue-Lafitte et al. revealed high MUC5AC expression in ulcerative colitis and to a lesser extent diverticulitis (50). Mucin-secreting cells in the colon express MUC5AC in response to bacterial and parasitic infections (51) and after perforation of the appendix associated with bacterial invasion (52). In the progression of colon polyps to cancer, Bu (45) and Kocer (42) both demonstrated that most conventional colorectal adenocarcinomas are negative for MUC5AC but most mucinous cancers are positive. MUC5AC RNA is known to be expressed to very high levels in serrated lesions of the colon (53,54), however two previous studies using differential transcriptomics did not reveal MUC5AC to have high sensitivity or specificity for SSLs (55,56). Therefore, we conclude that MUC5AC is normally expressed by mucous-secreting cells of the stomach, endocervis, gallbladder, and tracheobronchial tree but can be found in the large intestine in states of chronic inflammation, infection, and cancer in addition to pre-cancerous colon polyps.

## Discussion

Here, our search of mucin biomarkers reveals that MUC5AC differentiates serrated lesions of the colon from normal tissue. Further, the odds ratios suggest the expression rate of MUC5AC is SSL>HP>TSA>TA. This difference suggests there is a quantitative difference of MUC5AC expression among phenotypes, which may be more specific to SSLs. For example, Mikhaleva and colleagues report that MUC5AC expression is found intensely across the entire length of the crypt, while in HPs and TSAs expression is focal (57). Differences in MUC5AC expression between SSLs and other polyp types align with previous research on this topic. MUC5AC, MUC6, and MUC17 are consistently upregulated on SSLs based on evidence from transcriptomic, proteomic, and immunohistochemical studies.

Our findings are consistent with the notion that polyp subtypes, through differing carcinogenesis pathways, lead to significant differences in MUC5AC expression. The mechanism by which MUC5AC is upregulated is postulated to be through promoter hypomethylation and overexpression (Renault). DNA methylation changes has been found to be highly associated with serrated pathway lesions (32), and is correlated with BRAF mutation, CIMP, and MSI-H chromosomal phenotypes (32,54). In addition to alterations of mucin glycoproteins normally found in the colon, previous immunologic studies have shown that gastric antigens are expressed *de novo* in colonic polyps as well (58–60).

Other biomarkers for SSL development include Annexin A10 (61), Hes1 (62), Trefoil factor 1 (63), and Agrin in the muscularis layer (64), however few follow-up studies have validated these proteins. Of note, trefoil factors (TFFs) are often associated with mucins (65). Gastric-type mucin MUC6 was previously shown to be a promising biomarker for SSLs (34,38) but in a validation study did not demonstrate specificity (40), supporting the notion that HPs and SSLs lie on a continuum of carcinogenesis (54).

Although SSLs and TSAs arise from the serrated pathway, SSLs are recognized as a type of precursor lesion for colorectal cancer (CRC) with a microsatellite instability (MSI)-high phenotype, whereas TSA is likely to progress into CRC with a microsatellite stable (MSS) phenotype (66).

Given its sensitivity, we hypothesize that MUC5AC could be quantified on the surface of polyps and serve as a marker of SSL carcinogenesis. The difference in expression may distinguish HPs from SSLs for pathologists which is an evolving area of investigation. More broadly, akin to fecal immunochemical testing, the goal of a screening test is to identify as many individuals at-risk of a disease as possible. In the proper clinical context (i.e. excluding inflammatory bowel diseases and colitis patients), MUC5AC may fit this role to identify patients with SSL features. Further studies into the clinical utility of MUC5AC as a fecal biomarker or surface level expression are thus needed.

Our study has a few important limitations. Because of the difficulty in distinguishing SSLs from HPs, reported reclassification rates of HPs to SSLs in the 2007-2019 literature range from 2.6– 85%, which may explain why we found no significant difference in MUC5AC expression between SSLs and HPs. Thus, there is likely pathological overlap between the defined SSLs and HPs in the studies we analyzed. Technical factors, such as staining protocols and antibodies used, different definitions of thresholds to determine positivity, as well as possible selection bias with respect to the analyzed tumors may have caused these discrepancies.

## Data Availability

All data produced in the present work are contained in the manuscript

